# Public adherence to governmental recommendations regarding quarantine and testing for COVID-19 in two Norwegian cohorts

**DOI:** 10.1101/2020.12.18.20248405

**Authors:** Ellen Ø Carlsen, Ida Henriette Caspersen, Lill Trogstad, Håkon K Gjessing, Per Magnus

## Abstract

**Background:** Combatting the COVID-19 pandemic relies at present on non-pharmacological interventions. Governments are using various approaches from general advice to full lockdown. There is a need to describe and understand adherence to public health actions.

**Methods:** Participants from two ongoing cohorts, the Norwegian Mother, Father and Child Cohort Study (MoBa) and The Norwegian Influenza Pregnancy Cohort (NorFlu), answered questionnaires every 14 days since March 2020. From the summer of 2020, testing for presence of SARS-CoV-2 became easily available. Recommendations were that respiratory symptoms should lead to testing, and that confirmed or suspected COVID-19 should be followed by quarantine. We estimated the adherence to these guidelines in responses from cohort participants in the period August to October 2020.

**Results:** Less than 40% of men who were ill and less than 45% of women who were ill, tested themselves for SARS-CoV-2 during the same 14-day periods. Among subjects tested for COVID-19, about 53% of men and 59% of women reported quarantine. For subjects with a confirmed or suspected COVID-19 diagnosis, the proportions quarantined were 65% for men and 72% for women.

**Conclusions:** Public adherence to governmental recommendations regarding testing and quarantine were lower than expected in a country with high trust in government. This leaves considerable room for improvement in adherence, possibly reducing the need for more restrictive interventions.

## Introduction

Since COVID-19, a disease caused by severe acute respiratory syndrome coronavirus 2 (SARS-CoV-2), emerged in Wuhan, China, it has affected communities and societies worldwide. Epidemic modelling is useful in displaying scenarios under alternative and combined interventions. A recent model suggested that lockdowns may be necessary in the UK to prevent the health care system from breaking down.^1^ However, better empirical evidence is needed to corroborate predictions. Adherence to interventions is a central parameter in the models. Using responses from participants in two large, ongoing cohorts, our aim was to estimate the adherence to two governmental recommendations given to Norwegian citizens from August onwards: to test for SARS-CoV-2 when having respiratory symptoms, and to go into quarantine when a confirmed or suspected diagnosis of COVID-19 was made.

## Methods

The current study was based on The Norwegian Mother, Father and Child Cohort Study (MoBa)^2^ and The Norwegian Influenza Pregnancy Cohort (NorFlu),^3^ approved by The Regional Committees for Medical and Health Research Ethics, South East Norway C (no. 127708 for MoBa) and South East Norway B (no. 18403 for NorFlu).

Data collection in MoBa was licensed by the Norwegian Data Protection Agency and approved by The Regional Committees for Medical and Health Research Ethics. The data collection in the MoBa cohort is regulated by the Norwegian Health Registry Act.

### Participants

MoBa is an ongoing nationwide study in which more than 114,000 children and their parents, recruited during weeks 15-18 of gestation between 1999 and 2008, are followed up through questionnaires and linkages to registries. The participation rate was approximately 40%. During the swine flu pandemic (A H1N1(pdm09)) in 2009/2010, pregnant women were invited to participate in NorFlu which was established to examine associations between influenza and influenza vaccinations during pregnancy and a series of outcomes. About 9000 mothers and children are included. Both cohorts are followed by questionnaires, registry linkages and invitations into different sub-studies.

Sub-studies were initiated in both cohorts when the COVID-19 pandemic hit Norway. The participants have been invited to answer electronic questionnaires every 14 days with questions regarding illness, testing for COVID-19, quarantine, and more, starting in March/April 2020. About 150,000 adult participants were invited from MoBa as well as nearly 4500 women from NorFlu. Identical questionnaires have been used in the two cohorts. By November 1^st^, 16 questionnaires have been distributed. For MoBa, subjects who had not responded to the three first rounds in April were not invited to rounds 4 through 12, but again invited from round 13 onwards.

During the summer of 2020, the government recommended that all subjects who experience respiratory symptoms should be tested for presence of SARS-CoV-2. Thus, we included answers to questionnaire round numbers 11, 12, 13 and 14, covering calendar weeks 34 through 41, for analyses in the current study. The questionnaires were distributed in the period August 14^th^ – October 8^th^ for NorFlu and August 18^th^ - October 13^th^ for MoBa. The participation rates for rounds 11-14 for MoBa were 60% (reminders were not sent during summer), 83%, 55% and 58%, and for NorFlu 75%, 75%, 74% and 72%, respectively. The questionnaire information was linked to previous data from MoBa and NorFlu, to obtain information on educational level.

### Outcomes

The first outcome was being tested for SARS-CoV-2. The participants reported by answering *yes or no* to having been tested any time during *the previous 14 days*.

The second outcome was being in quarantine or self-isolation (asked in a single question, not differentiating between the two). The participants reported by answering *yes or no* to having been in quarantine or self-isolation any time during *the previous 14 days*.

### Exposures

The COVID-19 questionnaires provided information on the following exposures, as reported for the previous 14 days: 1) whether they had been ill with symptoms from the airways, had been feeling ill or had fever (in addition a series of specific symptoms were asked for); 2) had been tested for COVID-19; if tested, whether the test was positive (yes, no, do not know); 3) had been examined by or had a consultation with a physician and if yes, had received a diagnosis of confirmed or suspected COVID-19.

### Covariates

County of residence was obtained by linkage to the National Population Register for MoBa, but was not available for NorFlu participants. Information on educational level (less than high school, high school, university/college up to and including 4 years, more than 4 years of university/college) in the cohorts were available from previous questionnaire data. Age (displayed in 5- and 10-year intervals) and sex were derived from the previous cohort data.

### Statistical analysis

The proportions who reported to have been ill the previous 14 days, who had been tested for COVID-19, who tested positive among the tested (if answered “do not know” to the result of the test, we defined them as negative), who got a confirmed or suspected COVID-19 diagnosis by a physician, as well as the proportion who reported to have been in quarantine or self-isolation the previous 14 days, were examined for each questionnaire. To evaluate adherence to guidelines for testing over the study period (calendar weeks 34-41, 2020), we examined the proportion of tested people among those reporting illness the previous 14 days. We performed a sub-analysis by including only ill participants who reported at least one of the following symptoms: cough, dyspnoea, sore throat, and loss of taste or smell. Analyses stratified by gender, age group, questionnaire round, county of residence and educational level were performed. We included information from *the previous round* on having been ill with symptoms from the airways to examine whether this increased the adherence to testing. The association between illness and being tested for SARS-CoV-2 was assessed in logistic regression models, adjusting for round, age, educational level and county of residence, stratified by sex. To evaluate compliance with recommendations on quarantine/isolation, we examined the proportion who reported to have been in quarantine or isolation the previous 14 days according to: 1) having been tested for COVID-19, 2) having a positive COVID-19 test, 3) having a confirmed or suspected diagnosis of COVID-19 from a physician, 4) reporting illness. Proportions are reported according to round, county of residence, educational level and age, stratified by sex. To control for potential confounding, we ran logistic regression models adjusting for covariates.

All analyses were performed in Stata, (Statacorp, College Station, TX) version 16.0.

## Results

The proportion of participants who reported to have been ill the previous 14 days increased from 8.6% in round 11 to 13% in round 13, and then decreased again to 10% in round 14 (Table 1). The proportion tested for SARS-CoV-2 followed the same trend. Overall, the proportion that was tested among participants who had reported illness in the same 14-day period, was between 35% and 40% for men and between 40% and 45% for women (Figure 1). There was a tendency that the proportions were lowest for participants with highest education. Supplementary Table 1 gives the exact proportions for each round. The proportions increased slightly (2 to 5%) if we only included subjects with at least one of the following symptoms: cough, dyspnoea, sore throat, or loss of sense of taste or smell (Figure 1). We found no statistically significant differences in the proportions tested among men who fell ill when comparing age groups or educational levels. However, the odds ratio of being tested decreased in round 12 compared to round 11 and varied slightly by place of living in unadjusted and adjusted regression models (Supplementary Table 1). Among women, the odds ratios for being tested decreased in the later rounds, was higher for women aged 40-44 years compared to 45-49 years, and was lower among those with the highest educational level and varied by county of residence in both unadjusted and adjusted regression models (Supplementary Table 1). Inclusion of participants who had been ill with symptoms from the airways during *the previous round* lowered the proportion being tested (results not shown), suggesting that people are likely to get tested soon after the onset of symptoms.

**Table 1.** Characteristics of the study population.

|  | 14.08-31.08 | 28.08-15.09 | 11.09-29.09 | 25.09-13.10 |
| --- | --- | --- | --- | --- |
| **Characteristic** | **Round 11** | **Round 12** | **Round 13** | **Round 14** |
| **<u>ADULTS</u>** | **64318** | **91109** | **83795** | **88566** |
| **Age (years), n** |  |  |  |  |
| 25-34 | 563 | 762 | 699 | 711 |
| 35-39 | 5842 | 7846 | 7129 | 7521 |
| 40-44 | 18381 | 25651 | 23442 | 24828 |
| 45-49 | 23783 | 33775 | 31020 | 32810 |
| 50-54 | 12180 | 17636 | 16412 | 17305 |
| 55-59 | 2925 | 4366 | 4076 | 4313 |
| 60+ | 643 | 1070 | 1014 | 1075 |
| **Females, %** | 70 % | 63 % | 62 % | 63 % |
| **Ill last 14 days, %** | 8·6 % | 12 % | 13 % | 10% |
| **Tested for Covid last 14 days, %** | 5·3 % | 6·1 % | 6·9 % | 5·9 % |
| **Positive Covid test last 14 days among tested, % <sup>a</sup>** | 0·5 % | 0·2 % | 0·8 % | 0·8 % |
| **Confirmed or suspected Covid-diagnosis from doctor, %** | 0·7 % | 0·6 % | 0·5 % | 0·4 % |
| **Self-reported quarantine or isolation last 14 days, %** | 4·0 % | 4·7 % | 5·2 % | 4·2 % |

<sup>a</sup> those who answered "do not know" were set to negative

**Figure 1.**
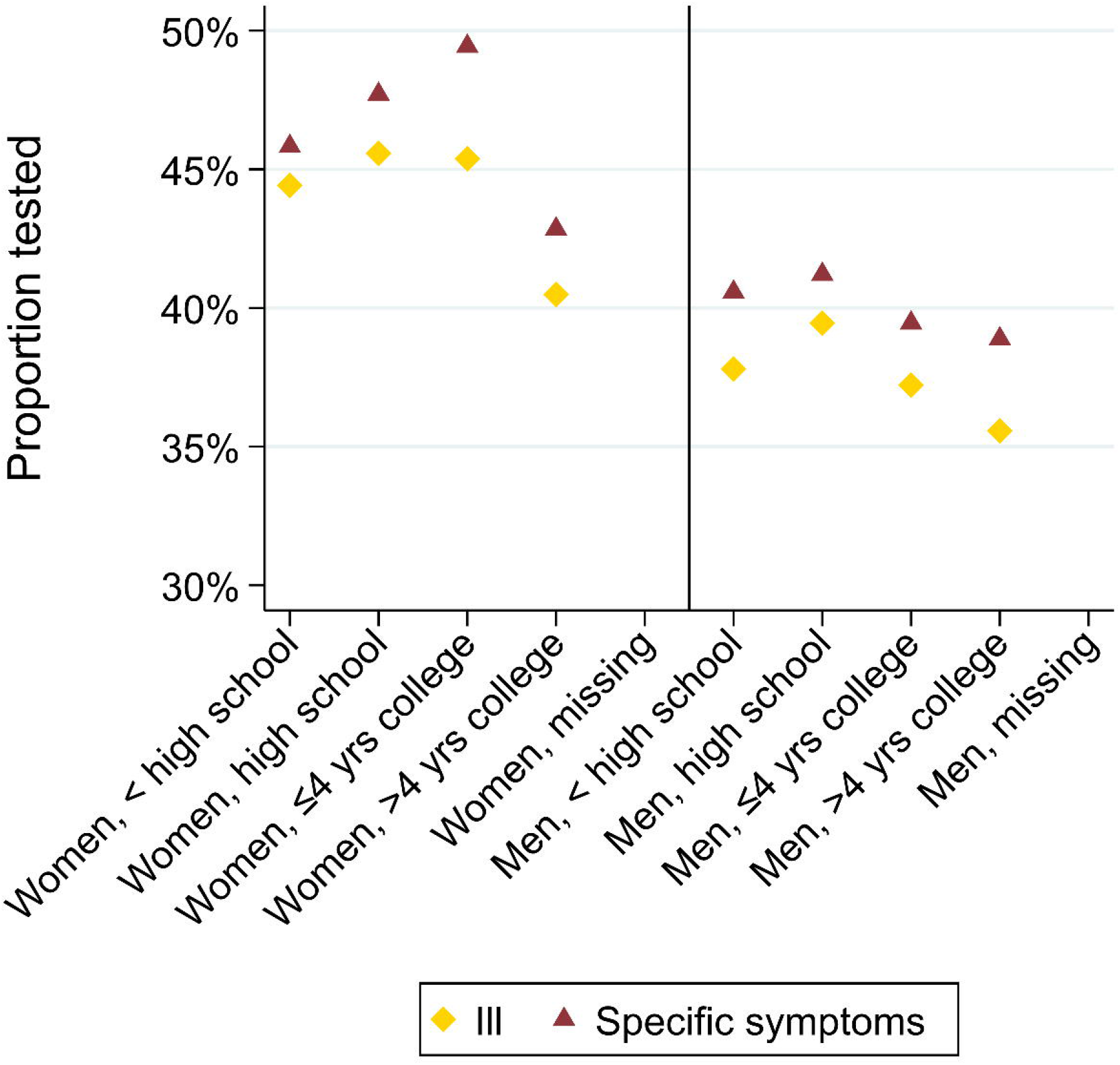
Proportions tested for COVID-19 last 14 days among those who report to have been ill the last 14 days, and among those who had at least one of the following symptoms: cough, dyspnoea, sore throat, fever or loss of taste/smell, stratified by sex and educational level. Answers from all the questionnaire rounds are included in the same analysis.

The proportion who reported to have been in quarantine or isolation was 4.0% in round 11, increased to 5.2% in round 13, and decreased to 4.2% in round 14 (Table 1). Among men who reported to have received a suspected or confirmed diagnosis of COVID-19 from a physician, 65% had been in quarantine or isolation during the same period, compared to 72% of the women, with slight fluctuations by educational level (Figure 2). The proportions of participants reporting quarantine or self-isolation were highest among subjects with a positive test for SARS-CoV-2, second highest among participants who received a diagnosis of confirmed or suspected COVID-19 from their physician, third highest among those who were tested for SARS-CoV-2, and lowest among those who were ill the previous 14 days (Figure 2, Supplementary Table 2). Among subjects tested for SARS-CoV-2, 53% of men and 59% of women reported to have been in quarantine or self-isolation. For respondents who had been ill during the previous 14 days, the numbers were 26% among men and 33% among women. Finally, 79% of men and 91% of women who reported having a positive SARS-CoV-2 test had followed the quarantine recommendations. The proportions who reported to have been in quarantine/self-isolation in different subgroups varied according to round in both crude and unadjusted analyses (Supplementary Table 2). County of residence also influenced the odds ratio of being in quarantine/self-isolation to a slight degree. For both women and men, age and educational level had a small effect on the odds ratio of quarantine/self-isolation among those who had been ill the previous 14 days and among those tested for SARS-CoV-2 (Supplementary Table 2).

**Figure 2.**
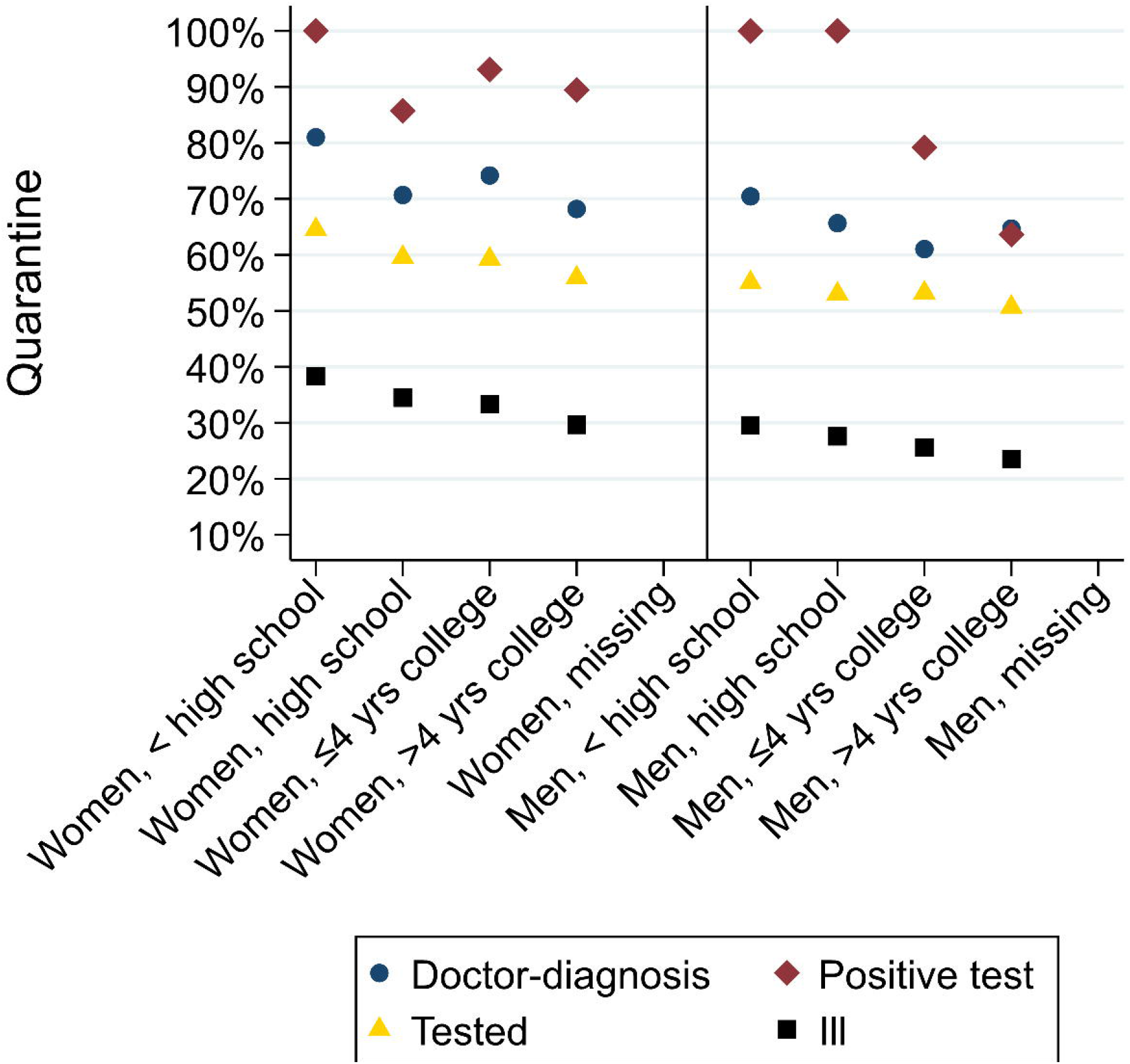
Proportion in quarantine/self-isolation among those who got a diagnosis of COVID-19 from their physician, tested positive for SARS-CoV-2, tested for SARS-CoV-2 regardless of result, and were ill. Stratified by sex and educational level. Answers from all the questionnaire rounds are included in the same analysis.

## Discussion

Using bi-weekly surveys, sent in August-October 2020 to ongoing population-based cohorts with an average of 85,000 respondents, we found relatively low adherence to government recommendations regarding testing for SARS-CoV-2 when ill, and to use of quarantine after receiving a diagnosis of suspected of confirmed COVID-19. The adherence was largely unaffected by age. The lowest adherence to the testing recommendations were among men and women with the highest education levels. Testing varied somewhat by county of residence, suggesting that local availability or guidelines may affect the decision to get tested.

To fight the pandemic, various restrictions and recommendations have been introduced, lifted, and re-introduced. In Norway, recommendations about quarantine were in place already from early March.^4^ From June 27^th^, testing was recommended for everyone with newly emerged symptoms of respiratory infection. Starting from August 12^th^ a strengthening of the recommendations included the following: anyone with symptoms of disease from the upper airways or other symptoms of COVID-19 should be tested.^5^ Furthermore, if tested due to symptoms, you should stay at home until the test-result is ready, and, regardless of testing, if you have symptoms of upper/lower airway disease, you should stay at home.^5,6^ Anyone with suspected or confirmed COVID-19 should be in quarantine/self-isolation.^6^ The following criteria for suspected COVID-19 were applied for the whole period under study: acute airway tract infection plus one of the following symptoms; fever, shortness of breath, cough, loss of sense of taste or smell, or examined by a physician who suspected COVID-19.^7^

Trust in government is high in Norway,^8,9^ suggesting that adherence may be even lower in other countries. Trust in the government and health care providers is also a predictor for using health care services.^10^ Public perceptions and opinions about governmental responses to the pandemic may certainly affect the level of acceptance of actions to mitigate the pandemic, and the government’s ability to adequately handle the crisis. Furthermore, people from higher socio-economic strata report a higher level of confidence in the government’s responses, although results varies across countries.^11^ As of October 27^th^, 1.6 million SARS-CoV-2 tests have been performed, and 18,341 confirmed cases and 280 deaths have been reported in Norway,^12^ which are relatively low numbers compared to other countries. We would thus assume a continued high trust in the government and high level of public perception of the government’s ability to handle the pandemic.^11^ However, a previous study among 1700 Norwegian participants responding to surveys from April to June, found low public adherence to quarantine/self-isolation.^13^ They found higher adherence among respondents required to be in quarantine/self-isolation who experienced symptoms compatible with COVID-19 (71%), while our results suggest that even among those who got tested for SARS-CoV-2, less than two thirds reported to have been in quarantine/self-isolation and even fewer among those with symptoms of illness the previous 14 days (less than one third) and in the sub-analysis including only specific symptoms.

One of the main strengths of our study is the continued high participation rates over more than seven months, particularly among the NorFlu cohort members. Both cohorts are population-based, and both sexes and all parts of the country are well represented in the study population. Using pre-existing cohorts as opposed to panels recruited during the pandemic has been outlined as advantageous in avoiding self-selection in recruitment during the pandemic.^14^ However, as the participants were originally recruited during pregnancy, all participants are parents and has an age distribution between 25 and 65+ years. Findings of willingness to adhere to governmental recommendations among adults may therefore not be generalizable to a childless population. Recruitment into the MoBa cohort has previously been studied, indicating a somewhat higher socio-economic status among participants than in the general Norwegian population.^15^ The same is likely to be true for the NorFlu cohort. Yet, we expect the proportion with respiratory illness/COVID-19 or COVID-like symptoms to be non-differentially distributed among cohort participants and non-participants of the same age groups in the general population. Also, immigrants to Norway are under-represented in the cohorts, and results from the current study cannot be generalized to this population. The relative proportion of COVID-19 in Norwegian residents with country background other than Norway has been higher than in residents with Norway as country of birth.^16^

As lifting of non-pharmacological interventions, such as banning of public events, have been associated with increased transmission rates,^17^ it seems vital that the public continues to follow the remaining recommendations from governments to be able to keep restrictions at a minimum. Self-isolation of symptomatic individuals has been stipulated to reduce transmission rates by 20-30%.^1^ Testing strategies and self-isolation or quarantine are recommendations that affect the public to a lesser degree than banning of public events, school closures, general requirements to stay at home or curfew. The implications on transmission rates of reduced public adherence to existing governmental recommendations need to be elucidated further both in other countries, and in settings with varying infection pressure. In this study, performed in a country with high trust in government, and a relatively low infection pressure during the study period, public adherence to governmental recommendations on regarding testing and quarantine were lower than expected. Accordingly, there is considerable room for improvement in adherence, possibly reducing the need for more restrictive interventions.

## Data Availability

The data underlying this article are available for analysis after approval from a Norwegian ethics committee and application to the Norwegian Institute of Public Health.

## Acknowledgments

The Norwegian Mother, Father and Child Cohort Study is supported by the Norwegian Ministry of Health and Care Services and the Ministry of Education and Research. We are grateful to all the participating families in Norway who take part in this on-going cohort study.

The Norwegian Influenza Pregnancy Cohort is supported by the Norwegian Institute of Public Health. We are deeply grateful to all the participating families for their continued and dedicated participation in the NorFlu cohort.

## Key-points

- Adherence to recommendations for testing and use of quarantine were lower than expected in a country with high trust in government
- Educational level, age, gender or county of residence did not influence the adherence to any large degree
- Our findings suggest that there is much to gain from stronger incentives for adhering to recommendations, which may alleviate the need for new lockdown situations

