## Supplementary Table 1 for "Public adherence to governmental recommendations regarding quarantine and testing for COVID-19 in two Norwegian cohorts"

**Supplementary Table 1.** Estimated odds ratios for getting tested for SARS-CoV-2 according to illness the previous 14 days. Stratified by sex, round, age, educational level, and county of residence.

| | Men | | | | Women | | | |
|---|---|---|---|---|---|---|---|---|
| | **Ill, n (%)** | **Tested among ill, n (%)** | **OR (95% CI) #, unadjusted** | **OR (95% CI) #, adjusted *** | **Ill, n (%)** | **Tested among ill, n (%)** | **OR (95% CI)#, unadjusted** | **OR (95% CI) #, adjusted *** |
| **Batch** | | | | | | | | |
| 11 | 1402 (7.3) | 560 (40.0) | 1 (Ref) | 1 (Ref) | 4106 (9.1) | 1886 (46.0) | 1 (Ref) | 1 (Ref) |
| 12 | 3202 (9.4) | 1127 (35.2) | 0.81 (0.72, 0.93) | 0.81 (0.71, 0.92) | 7808 (13.7) | 3250 (41.7) | 0.84 (0.78, 0.91) | 0.83 (0.77, 0.90) |
| 13 | 3250 (10.2) | 1208 (37.2) | 0.89 (0.78, 1.01) | 0.88 (0.77, 1.00) | 7481 (14.4) | 3296 (44.1) | 0.93 (0.86, 1.00) | 0.91 (0.84, 0.99) |
| 14 | 2822 (8.5) | 1076 (38.2) | 0.93 (0.81, 1.06) | 0.92 (0.80, 1.05) | 6362 (11.5) | 2789 (43.9) | 0.92 (0.85, 0.99) | 0.90 (0.82, 0.97) |
| **Age** | | | | | | | | |
| 25-34 | 43 (14.3) | 21 (48.8) | 1.63 (0.90, 3.0) | 1.53 (0.83, 2.80) | 475 (19.5) | 216 (45.6) | 1.13 (0.94, 1.37) | 1.03 (0.84, 1.26) |
| 35-39 | 714 (12.9) | 265 (37.1) | 1.01 (0.86, 1.19) | 0.98 (0.83, 1.15) | 3644 (16.0) | 1607 (44.2) | 1.07 (0.99, 1.16) | 0.97 (0.90, 1.06) |
| 40-44 | 2939 (10.9) | 1120 (38.1) | 1.06 (0.96, 1.16) | 1.04 (0.94, 1.15) | 9125 (14.0) | 4130 (45.3) | 1.12 (1.06, 1.19) | 1.12 (1.05, 1.19) |
| 45-49 | 4122 (9.3) | 1517 (36.9) | 1 (Ref) | 1 (Ref) | 8575 (11.2) | 3638 (42.5) | 1 (Ref) | 1 (Ref) |
| 50-54 | 2081 (7.4) | 756 (36.4) | 0.98 (0.88, 1.09) | 0.98 (0.88, 1.09) | 3441 (9.7) | 1424 (41.4) | 0.96 (0.89, 1.04) | 0.94 (0.87, 1.02) |
| 55-59 | 592 (6.3) | 221 (37.3) | 1.02 (0.85, 1.22) | 1.01 (0.84, 1.21) | 466 (7.5) | 193 (41.5) | 0.96 (0.80, 1.16) | 0.96 (0.79, 1.16) |
| 60+ | 185 (5.4) | 71 (38.4) | 1.07 (0.79, 1.44) | 1.11 (0.81, 1.50) | 29 (7.7) | 13 (44.8) | 1.10 (0.53, 2.30) | 0.96 (0.46, 2.01) |
| **Educational level** | | | | | | | | |
| < High school | 574 (7.8) | 217 (37.8) | 1.03 (0.85, 1.23) | 1.03 (0.86, 1.24) | 1023 (10.8) | 454 (44.4) | 0.96 (0.85, 1.10) | 0.99 (0.87, 1.13) |
| High school | 2711 (7.9) | 1068 (39.5) | 1.10 (0.99, 1.22) | 1.09 (0.99, 1.21) | 4986 (11.0) | 2269 (45.6) | 1.01 (0.94, 1.08) | 1.01 (0.94, 1.08) |
| College ≤4 years | 3558 (9.3) | 1323 (37.2) | 1 (Ref) | 1 (Ref) | 9838 (12.4) | 4462 (45.4) | 1 (Ref) | 1 (Ref) |
| College >4 years | 3560 (10.0) | 1264 (35.6) | 0.93 (0.85, 1.03) | 0.93 (0.84, 1.02) | 8501 (13.3) | 3441 (40.5) | 0.82 (0.77, 0.87) | 0.83 (0.78, 0.89) |
| Missing | 273 (10.2) | 99 (36.3) | 0.96 (0.74, 1.24) | 0.95 (0.73, 1.23) | 1409 (12.8) | 595 (42.3) | 0.88 (0.79, 0.99) | 0.77 (0.65, 0.90) |
| **County** | | | | | | | | |
| Oslo | 1331 (11.8) | 482 (36.4) | 1.03 (0.90, 1.18) | 0.37 (0.93, 1.22) | 2298 (14.3) | 921 (40.1) | 0.98 (0.89, 1.08) | 1.03 (0.93, 1.13) |
| Viken | 3170 (8.6) | 1128 (35.6) | 1 (Ref) | 1 (Ref) | 6889 (12.3) | 2785 (40.5) | 1 (Ref) | 1 (Ref) |
| Innlandet | 535 (7.4) | 196 (36.6) | 1.05 (0.86, 1.27) | 1.04 (0.86, 1.26) | 1397 (11.3) | 636 (45.6) | 1.23 (1.10, 1.39) | 1.22 (1.09, 1.37) |
| Vestfold&Telemark | 530 (9.2) | 218 (41.2) | 1.27 (1.05, 1.53) | 1.26 (1.05, 1.52) | 1147 (12.4) | 545 (47.6) | 1.34 (1.18, 1.52) | 1.32 (1.17, 1.50) |
| Agder | 596 (8.8) | 202 (34.0) | 0.93 (0.77, 1.12) | 0.93 (0.77, 1.11) | 1355 (12.0) | 556 (41.1) | 1.03 (0.91, 1.16) | 1.02 (0.91, 1.15) |
| Rogaland | 1032 (8.9) | 413 (40.1) | 1.21 (1.05, 1.40) | 1.20 (1.03, 1.38) | 2121 (11.9) | 997 (47.0) | 1.31 (1.19, 1.44) | 1.28 (1.16, 1.42) |
| Vestland | 1538 (9.7) | 607 (39.5) | 1.18 (1.04, 1.34) | 1.18 (1.04, 1.34) | 3953 (12.2) | 1919 (48.6) | 1.39 (1.29, 1.51) | 1.40 (1.29, 1.52) |
| Møre&Romsdal | 569 (8.4) | 218 (38.4) | 1.13 (0.94, 1.35) | 1.10 (0.91, 1.32) | 1433 (11.2) | 590 (41.2) | 1.03 (0.92, 1.16) | 1.01 (0.90, 1.14) |
| Trøndelag | 862 (8.9) | 335 (38.9) | 1.15 (0.99, 1.35) | 1.14 (0.98, 1.34) | 1994 (11.8) | 953 (47.8) | 1.35 (1.22, 1.49) | 1.34 (1.21, 1.49) |
| Nord-Norge | 507 (8.0) | 171 (33.7) | 0.92 (0.76, 1.12) | 0.91 (0.74, 1.10) | 1298 (11.5) | 496 (38.2) | 0.91 (0.81, 1.03) | 0.90 (0.80, 1.02) |

* Adjusted for round, educational level, county of residence and age.
### OR; Odds ratio, CI; Confidence interval
