## Supplementary Table 2 for "Public adherence to governmental recommendations regarding quarantine and testing for COVID-19 in two Norwegian cohorts"

**Supplementary Table 2**: Estimated odds ratios for quarantine/self-isolation according to the following exposures: 1) testing for SARS-CoV-2, 2) positive SARS-CoV-2 test, 3) suspected or confirmed diagnosis of COVID-19 from physician, and 4) ill the previous 14 days. Stratified by sex, round, county, age and educational level.

| | **Tested for Covid-19, n (%)** | **Quarantine among tested, n (%)** | **OR (95% CI) quarantine, unadjusted** | **OR (95% CI) quarantine, adjusted *** | **Pos Covid-test, n (% among tested)** | **Quarantine among positive, n (%)** | **Doctor-diagnosis Covid, n (%)** | **Quarantine among diagnosed, n (%)** | **OR (95% CI) quarantine, unadjusted** | **OR (95% CI) quarantine, adjusted *** | **Ill previous 14 days, n (%)** | **Quarantine among ill, n (%)** | **OR (95% CI) quarantine, unadjusted** | **OR (95% CI) quarantine, adjusted *** |
|---|---|---|---|---|---|---|---|---|---|---|---|---|---|---|
| **Sex** | | | | | | | | | | | | | | |
| Male | 5960 (5.1) | 3130 (52.5) | 1 (Ref) | 1 (Ref) | 52 (0.9) | 41 (78.9) | 501 (0.4) | 326 (65.1) | 1 (Ref) | 1 (Ref) | 10676 (9.0) | 2739 (25.7) | 1 (Ref) | 1 (Ref) |
| Female | 13946 (6.7) | 8147 (58.5) | 1.27 (1.20, 1.35) | 1.28 (1.20, 1.36) | 64 (0.5) | 58 (90.6) | 1222 (0.6) | 879 (72.1) | 1.39 (1.11, 1.73) | 1.37 (1.09, 1.71) | 25757 (12.3) | 8391 (32.6) | 1.40 (1.33, 1.47) | 1.39 (1.32, 1.46) |
| **Round**[§] | | | | | | | | | | | | | | |
| 11 | 3412 (5.3) | 1828 (53.6) | 1 (Ref) | 1 (Ref) | 16 (0.5) | 12 (75.0) | 429 (0.7) | 283 (66.1) | 1 (Ref) | 1 (Ref) | 5508 (8.6) | 1849 (33.6) | 1 (Ref) | 1 (Ref) |
| 12 | 5518 (6.1) | 3210 (58.3) | 1.21 (1.11, 1.31) | 1.23 (1.12, 1.34) | 10 (0.2) | 10 (100) | 525 (0.6) | 377 (71.8) | 1.31 (0.99, 1.72) | 1.38 (1.04, 1.84) | 11010 (12.1) | 3350 (30.5) | 0.87 (0.81, 0.93) | 0.87 (0.81, 0.93) |
| 13 | 5745 (6.9) | 3339 (58.2) | 1.20 (1.11, 1.31) | 1.22 (1.12, 1.34) | 46 (0.8) | 41 (89.1) | 405 (0.5) | 290 (72.0) | 1.31 (0.98, 1.77) | 1.39 (1.02, 1.89) | 10731 (12.8) | 3300 (30.8) | 0.88 (0.82, 0.94) | 0.88 (0.82, 0.94) |
| 14 | 5231 (5.9) | 2900 (55.5) | 1.08 (0.99, 1.18) | 1.10 (1.01, 1.20) | 44 (0.8) | 36 (81.8) | 364 (0.4) | 255 (70.1) | 1.20 (0.89, 1.62) | 1.23 (0.90, 1.68) | 9184 (10.4) | 2631 (28.7) | 0.80 (0.74, 0.85) | 0.79 (0.74, 0.85) |
| **County** | | | | | | | | | | | | | | |
| Oslo | 1985 (7.3) | 1036 (52.2) | 0.82 (0.73, 0.90) | 0.82 (0.74, 0.91) | 18 (0.9) | 18 (100) | 167 (0.6) | 112 (67.1) | 1.01 (0.70, 1.46) | 1.02 (0.70, 1.84) | 3629 (13.3) | 1033 (28.5) | 0.98 (0.90, 1.06) | 0.99 (0.91, 1.08) |
| Viken | 5210 (5.6) | 2980 (57.3) | 1 (Ref) | 1 (Ref) | 29 (0.6) | 24 (82.8) | 505 (0.5) | 337 (66.9) | 1 (Ref) | 1 (Ref) | 10059 (10.8) | 2907 (28.9) | 1 (Ref) | 1 (Ref) |
| Innlandet | 1044 (5.3) | 640 (61.4) | 1.19 (1.04, 1.36) | 1.17 (1.02, 1.34) | 3 (0.3) | 3 (100) | 100 (0.5) | 78 (78.8) | 1.84 (1.10, 3.08) | 1.85 (1.10, 3.11) | 1932 (9.9) | 609 (31.6) | 1.13 (1.02, 1.26) | 1.12 (1.01, 1.25) |
| Vestfold&Telemark | 943 (6.3) | 539 (57.2) | 1.00 (0.87, 1.15) | 1.00 (0.87, 1.15) | 2 (0.2) | 2 (100) | 79 (0.5) | 57 (72.2) | 1.28 (0.76, 2.17) | 1.30 (0.77, 2.01) | 1677 (11.2) | 537 (32.1) | 1.16 (1.04, 1.29) | 1.17 (1.04, 1.30) |
| Agder | 944 (5.2) | 476 (50.5) | 0.76 (0.66, 0.87) | 0.75 (0.66, 0.87) | 8 (0.9) | 5 (62.5) | 122 (0.7) | 81 (66.9) | 1.00 (0.66, 1.53) | 0.98 (0.64, 1.49) | 1951 (10.8) | 480 (24.6) | 0.80 (0.72, 0.90) | 0.80 (0.72, 0.90) |
| Rogaland | 1846 (6.3) | 1177 (63.8) | 1.32 (1.18, 1.47) | 1.33 (1.19, 1.48) | 4 (0.2) | 4 (100) | 172 (0.6) | 126 (73.3) | 1.36 (0.92, 2.00) | 1.36 (0.93, 2.01) | 3153 (10.7) | 1173 (37.2) | 1.46 (1.34, 1.58) | 1.48 (1.36, 1.61) |
| Vestland | 3494 (7.3) | 1925 (55.2) | 0.92 (0.84, 1.00) | 0.90 (0.83, 0.98) | 40 (1.2) | 37 (92.5) | 258 (0.5) | 189 (73.3) | 1.36 (0.97, 1.89) | 1.29 (0.92, 1.80) | 5491 (11.4) | 1799 (32.8) | 1.20 (1.12, 1.29) | 1.19 (1.11, 1.28) |

| | | | | | | | | | | | | | |
|---|---|---|---|---|---|---|---|---|---|---|---|---|---|
| Møre&Romsdal | 1049 (5.4) | 605 (57.8) | 1.02 (0.89, 1.17) | 1.02 (0.89, 1.17) | 3 (0.3) | 0 (0) | 85 (0.4) | 59 (69.4) | 1.12 (0.68, 1.85) | 1.11 (0.67, 1.82) | 2002 (10.2) | 588 (29.4) | 1.02 (0.92, 1.14) | 1.01 (0.91, 1.13) |
| Trøndelag | 1539 (5.8) | 851 (55.3) | 0.92 (0.82, 1.04) | 0.92 (0.82, 1.03) | 4 (0.3) | 3 (75.0) | 100 (0.4) | 77 (77.0) | 1.66 (1.00, 2.74) | 1.59 (0.96, 2.64) | 2856 (10.8) | 870 (30.5) | 1.08 (0.98, 1.18) | 1.07 (0.97, 1.17) |
| Nord-Norge | 861 (4.9) | 469 (54.6) | 0.90 (0.78, 1.04) | 0.89 (0.77, 1.03) | 4 (0.5) | 2 (50.0) | 54 (0.3) | 32 (59.3) | 0.72 (0.41, 1.28) | 0.74 (0.41, 1.31) | 1805 (10.2) | 503 (27.9) | 0.95 (0.85, 1.06) | 0.93 (0.83, 1.04) |
| **MEN** | | | | | | | | | | | | | | |
| **Age** | | | | | | | | | | | | | | |
| 25-34 | 27 (9.0) | 18 (66.7) | 1.83 (0.82, 4.09) | 1.80 (0.80, 4.04) | 1 (3.7) | 1 (100) | 5 (1.7) | 3 (60.0) | 0.79 (0.13, 4.86) | 0.63 (0.09, 4.43) | 43 (14.3) | 15 (34.9) | 1.52 (0.81, 2.86) | 1.34 (0.71, 2.52) |
| 35-39 | 361 (6.5) | 216 (59.8) | 1.36 (1.09, 1.71) | 1.34 (1.06, 1.68) | 1 (0.3) | 1 (100) | 44 (0.8) | 31 (7.5) | 1.26 (0.62, 2.57) | 1.10 (0.51, 2.36) | 714 (12.9) | 190 (26.6) | 1.03 (0.86, 1.23) | 0.97 (0.81, 1.17) |
| 40-44 | 1570 (5.8) | 865 (55.1) | 1.12 (0.99, 1.28) | 1.11 (0.97, 1.26) | 12 (0.8) | 10 (83.3) | 121 (0.5) | 81 (66.9) | 1.07 (0.66, 1.74) | 0.96 (0.58, 1.60) | 2939 (10.9) | 780 (26.6) | 1.03 (0.92, 1.15) | 1.01 (0.90, 1.12) |
| 45-49 | 2240 (5.0) | 1169 (52.2) | 1 (Ref) | 1 (Ref) | 16 (0.7) | 11 (68.8) | 188 (0.4) | 123 (65.4) | 1 (Ref) | 1 (Ref) | 4122 (9.3) | 1071 (26.0) | 1 (Ref) | 1 (Ref) |
| 50-54 | 1210 (4.3) | 626 (51.8) | 0.98 (0.85, 1.13) | 0.98 (0.85, 1.13) | 14 (1.2) | 11 (78.6) | 99 (0.4) | 63 (63.6) | 0.92 (0.56, 1.54) | 0.88 (0.52, 1.48) | 2081 (7.4) | 506 (24.3) | 0.91 (0.81, 1.03) | 0.91 (0.80, 1.02) |
| 55-59 | 407 (4.3) | 189 (46.4) | 0.79 (0.64, 0.98) | 0.78 (0.63, 0.96) | 6 (1.5) | 6 (100) | 36 (0.4) | 21 (58.3) | 0.74 (0.36, 1.53) | 0.83 (0.39, 1.77) | 592 (6.3) | 141 (23.9) | 0.89 (0.73, 1.09) | 0.86 (0.71, 1.06) |
| 60+ | 145 (4.3) | 47 (32.4) | 0.44 (0.31, 0.63) | 0.44 (0.30, 0.63) | 2 (1.4) | 1 (50.0) | 8 (0.2) | 4 (50.0) | 0.53 (0.13, 2.18) | 0.26 (0.05, 1.23) | 185 (5.4) | 36 (19.5) | 0.69 (0.47, 1.00) | 0.70 (0.48, 1.01) |
| **Educational level** | | | | | | | | | | | | | | |
| < High school | 369 (5.0) | 202 (55.0) | 1.08 (0.86, 1.35) | 1.10 (0.87, 1.38) | 5 (1.4) | 5 (100) | 44 (0.6) | 31 (70.5) | 1.52 (0.74, 3.14) | 1.76 (0.80, 3.87) | 574 (7.8) | 169 (29.6) | 1.22 (1.00, 1.48) | 1.23 (1.01, 1.49) |
| High school | 1658 (4.8) | 879 (53.0) | 0.99 (0.87, 1.13) | 0.94 (0.82, 1.08) | 8 (0.5) | 8 (100) | 166 (0.5) | 109 (65.7) | 1.22 (0.77, 1.92) | 1.30 (0.90, 2.10) | 2711 (7.9) | 747 (27.6) | 1.11 (0.99, 1.24) | 1.10 (0.98, 1.23) |
| College ≤4 years | 1942 (5.1) | 1033 (53.2) | 1 (Ref) | 1 (Ref) | 24 (1.2) | 19 (79.2) | 154 (0.4) | 94 (61.0) | 1 (Ref) | 1 (Ref) | 3558 (9.3) | 911 (25.6) | 1 (Ref) | 1 (Ref) |
| College >4 years | 1843 (5.2) | 934 (50.7) | 0.90 (0.79, 1.03) | 0.92 (0.81, 1.05) | 11 (0.6) | 7 (63.6) | 119 (0.3) | 77 (64.7) | 1.17 (0.71, 1.92) | 1.25 (0.74, 2.09) | 3560 (10.0) | 838 (23.6) | 0.89 (0.80, 1.00) | 0.90 (0.80, 1.00) |
| Missing | 148 (5.5) | 82 (55.4) | 1.09 (0.78, 1.53) | 1.09 (0.77, 1.53) | 4 (2.7) | 2 (50.0) | 18 (0.7) | 15 (83.3) | 3.19 (0.89, 11.5) | 4.79 (1.14, 20.1) | 273 (10.2) | 74 (27.1) | 1.08 (0.82, 1.42) | 1.08 (0.82, 1.43) |
| **WOMEN** | | | | | | | | | | | | | | |
| **Age** | | | | | | | | | | | | | | |
| 25-34 | 244 (10.0) | 170 (69.7) | 1.70 (1.28, 2.24) | 1.49 (1.10, 2.03) | 1 (0.4) | 1 (100) | 47 (1.9) | 33 (70.2) | 1.02 (0.53, 1.97) | 1.37 (0.61, 3.08) | 475 (19.5) | 201 (42.3) | 1.61 (1.33, 1.94) | 1.44 (1.17, 1.77) |
| 35-39 | 1933 (8.5) | 1183 (61.3) | 1.17 (1.05, 1.30) | 1.12 (1.00, 1.26) | 7 (0.4) | 7 (100) | 188 (0.8) | 139 (74.3) | 1.25 (0.85, 1.85) | 1.32 (0.86, 2.02) | 3644 (16.0) | 1254 (34.5) | 1.15 (1.06, 1.25) | 1.07 (0.98, 1.17) |
| 40-44 | 5001 (7.7) | 2959 (59.3) | 1.07 (0.99, 1.16) | 1.06 (0.98, 1.16) | 25 (0.5) | 21 (84.0) | 417 (0.6) | 313 (75.2) | 1.31 (0.96, 1.79) | 1.42 (1.03, 1.97) | 9125 (14.0) | 3092 (33.9) | 1.12 (1.06, 1.20) | 1.12 (1.05, 1.20) |
| 45-49 | 4574 (6.0) | 2626 (57.5) | 1 (Ref) | 1 (Ref) | 22 (0.5) | 21 (95.5) | 401 (0.5) | 280 (69.8) | 1 (Ref) | 1 (Ref) | 8575 (11.2) | 2683 (31.3) | 1 (Ref) | 1 (Ref) |

| | | | | | | | | | | | | | | |
|---|---|---|---|---|---|---|---|---|---|---|---|---|---|---|
| 50-54 | 1894 (5.4) | 1055 (55.8) | 0.93 (0.84, 1.04) | 0.94 (0.84, 1.05) | 7 (0.4) | 6 (85.7) | 142 (0.4) | 100 (70.9) | 1.05 (0.69, 1.61) | 1.08 (0.70, 1.67) | 3441 (9.7) | 1017 (29.6) | 0.92 (0.85, 1.00) | 0.92 (0.84, 1.00) |
| 55-59 | 275 (4.5) | 139 (50.7) | 0.76 (0.60, 0.97) | 0.75 (0.58, 0.96) | 0 (0) | 0 (0) | 22 (0.4) | 13 (59.1) | 0.62 (0.26, 1.50) | 0.60 (0.24, 1.47) | 466 (7.5) | 132 (28.5) | 0.87 (0.71, 1.07) | 0.85 (0.69, 1.05) |
| 60+ | 24 (6.4) | 15 (65.2) | 1.38 (0.59, 3.27) | 1.43 (0.60, 3.40) | 2 (8.3) | 2 (100) | 5 (1.3) | 1 (20.0) | 0.11 (0.01, 0.98) | 0.09 (0.01, 0.79) | 29 (7.7) | 12 (41.4) | 1.55 (0.74, 3.24) | 1.46 (0.70, 3.08) |
| **Educational level** | | | | | | | | | | | | | | |
| < High school | 600 (6.3) | 386 (64.6) | 1.25 (1.05, 1.49) | 1.22 (1.02, 1.46) | 1 (0.2) | 1 (100) | 79 (0.8) | 64 (81.0) | 1.48 (0.82, 2.70) | 1.51 (0.82, 2.80) | 1023 (10.8) | 391 (38.3) | 1.24 (1.09, 1.42) | 1.23 (1.07, 1.41) |
| High school | 2881 (6.4) | 1714 (59.6) | 1.01 (0.92, 1.11) | 1.00 (0.90, 1.09) | 14 (0.5) | 12 (85.7) | 332 (0.7) | 234 (70.7) | 0.84 (0.61, 1.15) | 0.83 (0.60, 1.16) | 4986 (11.0) | 1717 (34.5) | 1.05 (0.98, 1.13) | 1.04 (0.97, 1.12) |
| College ≤4 years | 5410 (6.8) | 3198 (59.3) | 1 (Ref) | 1 (Ref) | 29 (0.5) | 27 (93.1) | 463 (0.6) | 342 (74.2) | 1 (Ref) | 1 (Ref) | 9838 (12.4) | 3273 (33.3) | 1 (Ref) | 1 (Ref) |
| College >4 years | 4325 (6.8) | 2416 (55.9) | 0.87 (0.80, 0.94) | 0.90 (0.82, 0.98) | 19 (0.4) | 17 (89.5) | 283 (0.4) | 193 (68.2) | 0.75 (0.54, 1.03) | 0.80 (0.57, 1.13) | 8501 (13.3) | 2518 (29.7) | 0.84 (0.79, 0.90) | 0.86 (0.80, 0.92) |
| Missing | 730 (6.6) | 433(59.5 ) | 1.01 (0.86, 1.18) | 1.04 (0.83, 1.20) | 1 (0.1) | 1 (100) | 65 (0.6) | 46 (70.8) | 0.84 (0.47, 1.50) | 0.92 (0.40, 2.10) | 1409 (12.8) | 492 (35.0) | 1.08 (0.96, 1.21) | 1.07 (0.91, 1.26) |

§ Round 11 14/8-31/8, round 12 28/8-15/9, round 13 11/9-29/9, round 14 25/9-13/10, 2020.
